# An individualized risk prediction model for new-onset, progression and regression of chronic kidney disease in a retrospective cohort of patients with type 2 diabetes under primary care in Hong Kong

**DOI:** 10.1101/2020.01.28.20019182

**Authors:** Lin Yang, Tsun Kit Chu, Jinxiao Lian, Cheuk Wai Lo, Shi Zhao, Daihai He, Jing Qin, Jun Liang

## Abstract

**Objectives:** This study is aimed to develop and validate a prediction model for multi-state transitions across different stages of chronic kidney disease in patients with type 2 diabetes mellitus under primary care.

**Setting:** We retrieved the anonymized electronic health records of a population based retrospective cohort in Hong Kong.

**Participants:** A total of 26,197 patients were included in the analysis.

**Primary and secondary outcome measures:** The new-onset, progression, and regression of chronic kidney disease were defined by the transitions of four stages that were classified by combining glomerular filtration rate and urine albumin-to-creatinine ratio. We applied a multi-scale multi-state Poisson regression model to estimate the rates of the stage transitions by integrating the baseline demographic characteristics, routine laboratory test results and clinical data from electronic health records.

**Results:** During the mean follow-up time of 1.7 years, there were 2,935 patients newly diagnosed with chronic kidney disease, 1,443 progressed to the next stage and 1,971 regressed into an earlier stage. The models achieved the best performance in predicting the new-onset and progression with the predictors of sex, age, body mass index, systolic blood pressure, diastolic blood pressure, serum creatinine, HbA1c, total cholesterol, LDL, HDL, triglycerides and drug prescriptions.

**Conclusions:** This study demonstrated that individual risks of new-onset and progression of chronic kidney disease can be predicted from the routine physical and laboratory test results. The individualized prediction curves developed from this study could potentially be applied to routine clinical practices, to facilitate clinical decision making, risk communications with patients and early interventions.

**Article summary:** *Strengths of this study:*

- Early predictions for chronic kidney disease progression and timely intervention is critical for clinical management of patients with diabetes.
- We successfully developed a multi-scale multi-state Poisson regression models that achieved the satisfactory performance in predicting the new-onset and progression of chronic kidney diseases.
- The model incorporates the predictors of demographic characteristics, routine laboratory test results and clinical data from electronic health records.
- The individualized prediction curves could potentially be applied to facilitate clinical decision making, risk communications with patients and early interventions of CKD progression.

*Limitations of this study:*

- The cohort has a relatively short follow-up period and the retrospective study design might suffer from report bias and selection bias.

## Introduction

Globally, in 2017 there were 425 million adults with diabetes according to the International Diabetes Federation ^1^. In Hong Kong, it was estimated that 10.9% of adults aged 20-79years had diabetes ^1^. Diabetes is associated with heavy disease burden and tremendous economic costs ^2^. A local study reported that the annual direct medical costs of diabetes patients with newly diagnosed complications were much higher than the cost of US$1,984 of uncomplicated cases ^3^. Chronic kidney disease (CKD) is one of the most common complications of type 2 diabetes. Globally, it was estimated that diabetes attributed to 12-55% of end-stage renal disease (ESRD) ^1^. In Hong Kong, a prospective cohort of the Hong Kong Diabetes Registry reported that 10% of type 2 diabetes patients developed into CKD, which was defined as estimated glomerular filtration rate (eGFR) < 60ml/min per 1.73m^2^, after seven years of follow-up ^4^.

Early diagnosis and treatment of CKD in patients with type 2 diabetes is critical for reducing its associated heavy disease burden. Therefore, developing an effective and efficient prediction model for CKD progression is important in term of prioritizing healthcare resources to high-risk populations. The KDIGO 2012 Guideline lists several predictors for CKD progression based on previous epidemiological studies, including age, sex, ethnicity, obesity, CKD etiology, GFR, albuminuria, blood pressure, glycaemic control, dyslipidemia, and complications of cardiovascular diseases ^5^. Specifically for people with type 2 diabetes, several prediction models have been reported in literature, but most were Cox proportional hazards regression models on all-cause mortality and incidence of cardiovascular disease ^6-9^. Previous studies have investigated the risk factors that significantly predicted ESRD in Chinese populations ^10 11^, but none focused on CKD progression at earlier stages nor on CKD regression.

The Cox proportional hazards model is one of the most widely adopted classical modeling approaches to predict diabetes complications. However, it only allows two states (no event and event) and linear predictors, hence it is unable to assess the progression probability across different disease stages. Some studies adopted more complex Cox models to address these limitations, such as multi-state Markov model and the Cox model with time-dependent covariates ^12 13^. But these models include only a single time scale, which usually does not allow for the simultaneous estimation on the forward and backward transitions of multiple disease stages (i.e. progression and regression).

A multi-scale multi-state Poisson regression (MSMS) model has recently been applied to estimate the risks of transition between different stages of disease progression or the incidences of multiple intermediate states and endpoints such as different hospitalization episodes ^14^. This model has the advantages of allowing multiple events in the follow-up period and thereby is capable of investigating dynamic transition across different stages of disease progression and regression. In other words, forward time scales (for example, time since enrolment into the cohort) and backward time scales (time since entry to the intermediate state) are simultaneously entered into the model as covariates. Another advantage of the MSMS model lies in its flexibility of allowing nonlinear dynamic transitions by incorporating spline smooth functions in each timescale with full parametric estimation. Here we conducted a population based retrospective cohort study, with the aim to develop and validate a prediction model for the new-onset, progression and regression of CKD in patients with type 2 diabetes under primary care in Hong Kong.

## Methods

### Data sources

We obtained individual data of adults who were aged over 18 years at enrolment into the Risk Assessment and Management Programme for Patients with Diabetes Mellitus (RAMP-DM) during July 2014 to June 2017, in the general and specialty outpatient clinics managed by the New Territory West Cluster of the Hospital Authority in Hong Kong Special Administrative Region, China. This cluster served a population of 1.1 million in 2017, and more details about the data source can be found in reference ^15^. The RAMP-DM program aims to structurally screen for diabetic complications among patients with diagnosed diabetes type 1 or type 2 under primary care. All patients are eligible to join this program without extra costs and on a voluntary basis. Doctors and nurses from different hospitals and clinics were regularly trained and followed the same protocol of RAMP-DM.

We also retrieved the longitudinal data of physical examinations and laboratory test results during scheduled clinical visits, including incidence of diabetic complications, blood pressure, eGFR (calculated from urine creatinine), urine albumin-to-creatinine ratio (UACR), total cholesterol, serum levels of creatinine, HDL, LDL, triglycerides and serum HbA1c, together with annual prescriptions of angiotensin-converting-enzyme inhibitors (ACEI) and angiotensin II receptor blockers (ARB), from the Clinical Data Analysis and Reporting System during 1 January 2014 to 31 December 2017, by matching their unique patient reference numbers. All adults aged above 18 years with clinical diagnosed type 2 diabetes with or without complications, according to the International Classification of Disease Ninth Revision codes (250.00, 250.02, 250.10, 250.12, 250.20, 250.22, 250.30, 250.32, 250.40, 250.42, 250.50, 250.52, 250.60, 250.62, 250.70, 250.72, 250.80, 250.82, 250.90, 250.92), were included in the analysis. The participants who already had ESRD at enrolment, without HbA1C tests taken during the follow-up, and or having incomplete baseline data were excluded from data analysis.

#### Patient and Public Involvement statement

Patients were not involved in the recruitment to and conduct of the study, since all data were retrospectively retrieved from electronic medical records.

#### Definition of CKD new-onset, progression and regression

According to the KDIGO 2012 Clinical Practice Guideline for Evaluation and Management of Chronic Kidney Diseases ^5^, CKD can be classified into four risk stages for CKD outcomes based on eGFR and UACR levels: stage 0, low risk; stage 1, moderately increased risk; stage 2, high risk; stage 3, very high risk. Here we defined three categories of stage transitions as event: 1) *new-onset of CKD*, transition from stage 0 to 1; 2) *CKD progression*, forward transition to the next advanced stage (i.e. stage 1 to 2, 2 to 3), and 3) *CKD regression*, backward transition to the next stage (i.e. stage 1 to 0, 2 to 1, and 3 to 2). Cases with transition over two stages with the intermediate stage recorded were counted as two separate events, and those without were excluded from the study. For example, two events were included in the model if the subject was classified into stage 0 at the beginning, and progressed into stage 1 and then stage 2 in the follow-up period. If a subject jumped from stage 0 to 2 without a record of stage 1, his/her data was not included in the model.

#### Data matching

The eGFR and UACR test results taken on the same date of CKD outcome events (new-onset, progression and regression) were matched by the unique patient numbers. If these tests were conducted on different dates, those taken closest to the event dates and within 180 days were matched to the CKD event dates with the unique patient numbers. Baseline sociodemographic data were collected at the initial visits to the clinics, including age, sex, education level, body mass index (BMI) and the presence of diabetic complications. Longitudinal data of blood pressure, lipid profile, serum creatinine, eGFR, UACR and HbA1c levels tested during the clinical visits, together with any ACEI/ARB prescriptions within the year (yes or no), were also matched for each participant by the dates of follow-up period between recruitment dates and event dates, or between recruitment dates and censoring dates if no events occurred by the end of follow-up period.

#### MSMS model

The technical details of MSMS models can be found in references ^14 16^. We used calendar time as the basic time scale, and the risk of transition was calculated for the period from the enrolment time to the lab test dates with stage change. The incidence rate of the new-onset of CKD (or progression, regression) was estimated from the following formula:

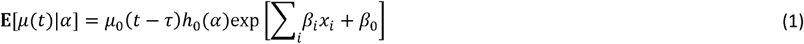

where *t* is the time since enrolment, E[*μ*(*t*)|α] is the expected crude incidence rate of CKD new-onset (or progression, regression) from the diagnosis of the previous stage to time *t*, which is assumed to follow a Poisson distribution; *τ* is the time when last lab tests are conducted for CKD (*t* > *τ* ≥ 0); *α* is the most recent HbA1c levels prior to the progression events; *x*_i_s are the covariates of age, sex, BMI, systolic blood pressure (SBP), diastolic blood pressure (DBP), serum creatinine, total cholesterol, HDL, LDL, triglycerides that were tested at enrolment, and annual drug prescriptions of ACEI and ARB; *μ*_*0*_(*t* − *τ*) is the natural splines function of the period (*t* − *τ*) during which the transition occurs; *h*_*0*_ (*α*) denotes the natural splines of HbA1c levels (*α*). Here we assume nonlinear effects of time and HbA1c level, because the incidence rate of CKD new-onset (or progression, regression) could probably increase over time in a nonlinear pattern. Unlike the model proposed by Lacobelli & Carstensen ^14^, we only included one time scale *t* − *τ* in this model to simplify the model structures, because there were very limited numbers of participants who experienced both the regression and progression events in the follow-up period.

We first randomly extracted 90% of the cohort data as a training dataset to fit the MSMS model by 9-fold cross-validation ^17^. The remaining 10% of data were used as the test dataset to evaluate the prediction accuracy of the best-fit model for internal validation. For each outcome, we built four different models: model 1 included sex, age, BMI and HbA1c as predictor; model 2, blood pressure and total cholesterol were added to model 1; model 3, serum creatinine, HDL, LDL and triglycerides were added to model 2; model 4, annual prescription of ACEI and ARB were added to model 3. These models with different combinations of predictors were compared using the area under the ROC curve (AUC) and mean absolute error (MAE) ^18 19^. The Nagelkerke scaled R^2^ was adopted to measure the goodness-of-fit of model ^20^.

Individual incidence rates of CKD new-onset, progression and regression over time since enrolment could be predicted from the final MSMS model by inputting the observed data of each subject. It is of note that the time between laboratory tests and clinical visits was not consistent, as they were determined by clinical indications and hospital manpower. Hence, to adjust for irregularly scheduled clinical visits and tests, we calculated an adjusted incidence rate by adding weights of the probability density function of clinical visits and test frequency, with the following equation 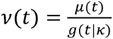, where *ν*(*t*) is the adjusted CKD incidence rate; *g*(*t*|*κ)* denotes the Gaussian kernel density function of clinical visits and tests. An example of this adjustment is shown in Appendix 1.

All the data analyses were conducted in R (version 3.4.1) and the codes can be found at https://github.com/yanglinpolyu/DMforecastmodel.

## Results

### Summary statistics

A flow chart for data collection and analysis is shown in Figure 1. There were 40,781 people with diabetes who were enrolled into the RAMP-DM from July 2014 to June 2017. Of them, 39,652 people with type 2 diabetes were considered eligible for this study. We excluded 8,078 patients with incomplete eGFR and UACR data, 5,002 with only one record of eGFR and UACR, and 375 with incomplete HbA1c record. In the final sample of 26,197 patients, 48.8% were men, the mean age at enrolment was 61.5 years, and 70.8% did not have CKD (stage 0). At the end of follow-up period, there were 2,632 cases of CKD new-onset, 1,746 progression, and 1,971 regression (Table 1). Compared to the patients without change, those with the new-onset or progression or regression of CKD were older, more likely to be female, with lower education level and higher BMI, having more complications (Table 2). The mean (range) of the follow-up period was 1.84 (0.07 - 4.04), 1.77 (0.13 - 3.84), 1.45 (0.08 - 3.72) and 1.89 (0.07 - 4.04) years, for all patients, patients with CKD new-onset/progression, with regression, and without change, respectively.

**Figure 1.**
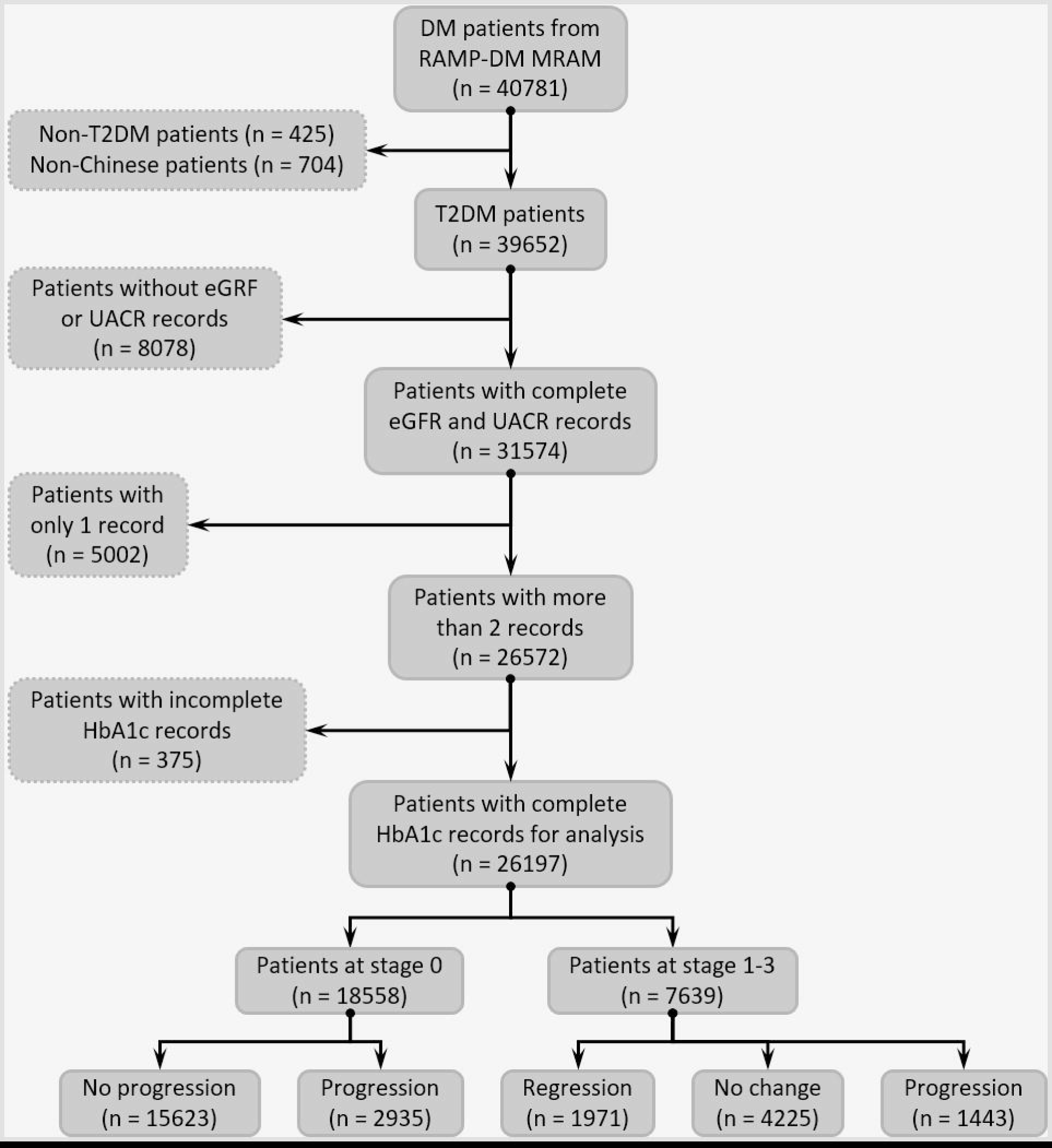
Flow chart of data clean and analysis procedure for CKD progression.

**Table 1.**
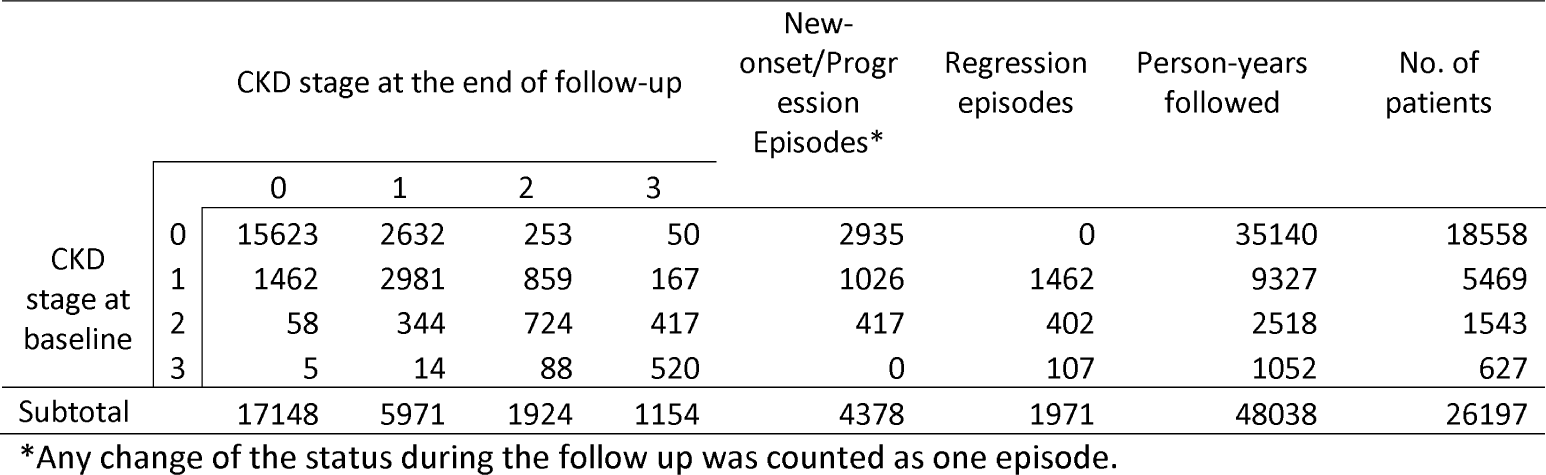
Summary of CKD progression and regression during the follow up period.

| CKD stage at baseline | CKD stage at the end of follow-up |  |  |  | New-onset/Progression Episodes* | Regression episodes | Person-years followed | No. of patients |
| --- | --- | --- | --- | --- | --- | --- | --- | --- |
|  | 0 | 1 | 2 | 3 |  |  |  |  |
| 0 | 15623 | 2632 | 253 | 50 | 2935 | 0 | 35140 | 18558 |
| 1 | 1462 | 2981 | 859 | 167 | 1026 | 1462 | 9327 | 5469 |
| 2 | 58 | 344 | 724 | 417 | 417 | 402 | 2518 | 1543 |
| 3 | 5 | 14 | 88 | 520 | 0 | 107 | 1052 | 627 |
| Subtotal | 17148 | 5971 | 1924 | 1154 | 4378 | 1971 | 48038 | 26197 |

\*Any change of the status during the follow up was counted as one episode.

**Table 2.** Descriptive statistics of cohort participants.

|  | Total | No change | Regression | New-onset/<br>Progression |
| --- | --- | --- | --- | --- |
| **n** | 26197 | 19848 | 1971 | 4378 |
| **Person-years** | 48038 | 37478 | 2817 | 7743 |
| **Sociodemographic factors** |  |  |  |  |
| Sex, n(%) |  |  |  |  |
| Female | 12965 (49.5%) | 9458 (47.7%) | 1138 (57.7%) | 2369 (54.1%) |
| Male | 13232 (50.5%) | 10390 (52.3%) | 833 (42.3%) | 2009 (45.9%) |
| Age, mean±sd | 62.4 ± 10.4 | 61.5 ± 10.1 | 64.0 ± 11.0 | 65.7 ± 10.8 |
| Education levels, n(%) |  |  |  |  |
| No formal education | 3150 (12.0%) | 2094 (10.6%) | 322 (16.3%) | 734 (16.8%) |
| Primary | 9861 (37.6%) | 7320 (36.9%) | 779 (39.5%) | 1762 (40.2%) |
| Secondary | 11734 (44.8%) | 9272 (46.7%) | 771 (39.1%) | 1691 (38.6%) |
| Tertiary | 1368 (5.2%) | 1095 (5.5%) | 92 (4.7%) | 181 (4.1%) |
| Other | 84 (0.3%) | 67 (0.3%) | 7 (0.4%) | 10 (0.2%) |
| BMI, mean±sd | 26.2 ± 4.2 | 26.1 ± 4.1 | 26.8 ± 4.2 | 26.4 ± 4.6 |

Table 2. Continued.
|  | Total | No change | Regression | New-onset/<br>Progression |
| --- | --- | --- | --- | --- |
| BMI category, n(%) |  |  |  |  |
| Underweight | 286 (1.1%) | 207 (1.0%) | 25 (1.3%) | 54 (1.2%) |
| Normal | 5180 (19.8%) | 4013 (20.2%) | 309 (15.7%) | 858 (19.6%) |
| Overweight | 5565 (21.3%) | 4333 (21.8%) | 379 (19.3%) | 853 (19.5%) |
| Obese | 15140 (57.9%) | 11280 (56.9%) | 1254 (63.8%) | 2606 (59.6%) |
| **Clinical characteristics** |  |  |  |  |
| Duration of diabetes (years, mean±sd) | 7.8 ± 6.3 | 7.5 ± 6.1 | 7.9 ± 6.2 | 9.2 ± 6.8 |
| Coronary heart disease, n(%) | 561 (2.1%) | 386 (2.1%) | 66 (3.3%) | 109 (2.5%) |
| Stroke, n(%) | 993 (3.8%) | 669 (3.4%) | 81 (4.1%) | 243 (5.6%) |
| Peripheral arterial disease, n(%) |  |  |  |  |
| Yes | 252 (1.0%) | 173 (0.9%) | 27 (1.4%) | 52 (1.2%) |
| Suspected | 117 (0.4%) | 87 (0.4%) | 5 (0.3%) | 25 (0.6%) |
| Diabetic retinopathy, n(%) |  |  |  |  |
| No retinopathy | 17388 (66.4%) | 13530 (68.2%) | 1259 (63.9%) | 2599 (59.4%) |
| Non-sight threatening | 5341 (20.4%) | 4014 (20.2%) | 409 (20.8%) | 918 (21.0%) |
| Sight threatening | 2651 (10.1%) | 1760 (8.9%) | 215 (10.9%) | 676 (15.4%) |
| Ungradable | 176 (0.7%) | 117 (0.6%) | 21 (1.1%) | 38 (0.9%) |
| SBP (mmHg, mean±sd) | 131.6 ± 16.3 | 131.1 ± 16.1 | 132.9 ± 17.1 | 133.5 ± 17.0 |
| DBP (mmHg, mean±sd) | 75.0 ± 10.3 | 75.2 ± 10.1 | 74.6 ± 10.9 | 74.1 ± 10.8 |
| **Laboratory tests, mean±sd** |  |  |  |  |
| UACR (mg/mmol) | 5.6 ± 21.4 | 5.5 ± 21.6 | 10.9 ± 21.0 | 6.0 ± 20.0 |
| eGFR (ml/min/1.73 m<sup>2</sup>) | 84.3 ± 19.5 | 86.5 ± 18.7 | 78.8 ± 21.7 | 77.1 ± 19.8 |
| Creatinine (mmol/L) | 79.2 ± 21.3 | 77.7 ± 20.5 | 83.5 ± 25.4 | 84.1 ± 21.7 |
| HbA1c (%) | 7.2 ± 1.3 | 7.1 ± 1.3 | 7.3 ± 1.3 | 7.2 ± 1.3 |
| HbA1c (mmol/mol) | 55.2 ± 11.9 | 54.1 ± 11.9 | 56.3 ± 11.9 | 55.2 ± 11.9 |
| Triglycerides (mmol/L) | 1.5 ± 1.1 | 1.5 ± 1.1 | 1.6 ± 1.2 | 1.6 ± 1.0 |
| LDL (mmol/L) | 2.3 ± 0.7 | 2.3 ± 0.7 | 2.4 ± 0.7 | 2.3 ± 0.7 |
| HDL (mmol/L) | 1.3 ± 0.3 | 1.3 ± 0.3 | 1.3 ± 0.3 | 1.2 ± 0.3 |
| **Drug prescriptions, n(%)** |  |  |  |  |
| ACEI | 11259 (43.0%) | 7992 (40.3%) | 991 (50.3%) | 2276 (52.0%) |
| ARB | 6964 (26.6%) | 4694 (23.6%) | 690 (35.0%) | 1580 (36.1%) |

Abbreviations: BMI, body mass index; SBP, systolic blood pressure; DBP, diastolic blood pressure; UACR, urine albumin-to-creatinine ratio; ACEI, angiotensin-converting-enzyme inhibitors; ARB, angiotensin II receptor blockers.

### Model performance and predictors

The MSMS model achieved better performance in predicting progression than in predicting the regression of chronic kidney disease (AUC 0.72-0.84 vs 0.50-0.71) (Appendix 2). The significant predictors for CKD new-onset included female, older age, having ACEI and ARB prescriptions, high levels of BMI, SBP, total cholesterol and serum creatinine, low levels of DBP, HDL and LDL (Appendix 3). Fewer significant predictors were found for those who progressed from stage 1 to 2 and from 2 to 3, and the effect estimates of significant predictors were similar to those of new-onset. The only exception is that lower BMI was associated with a higher risk of progression from stage 2 to 3.

### Rates of new-onset and progression

To facilitate the comparison between groups, we plotted the prediction curves of adjusted rates per 1,000 person-year by assuming the mean values of each predictor in the MSMS model: BMI = 25.7, SBP = 130mmHg, DBP = 75mmHg, total cholesterol = 4.0 mmol/l, Creatinine = 75 mmol/l, HDL = 1.20 mmol/l, LDL = 2.10 mmol/l, Triglycerides = 1.21 mmol/l. The mean age was 56.4 years for female aged <65yrs, 73.7 for female aged 65+yrs, 56.1 for male aged <65yrs, and 72.4 for male aged 65+yrs.

The temporal trend of adjusted rates of CKD new-onset and progression from the MSMS model across different levels of HbA1c are shown in Figure 2 and 3, for women and men, respectively. The CKD progression rate dramatically increased in women two years after diagnosis, but this change was less evident in men. An exponential increasing trend of incidence rates over time was observed for all three types of CKD progression, and gradually elevated when HbA1c level increased from 6% to 9%. The new-onset rate was high within the first year of enrolment (diagnosed as stage 0), particularly among older adults aged 65 years or over. The progression rate from stage 2 to 3 dramatically increased two years after the diagnosis of stage 2.

**Figure 2.**
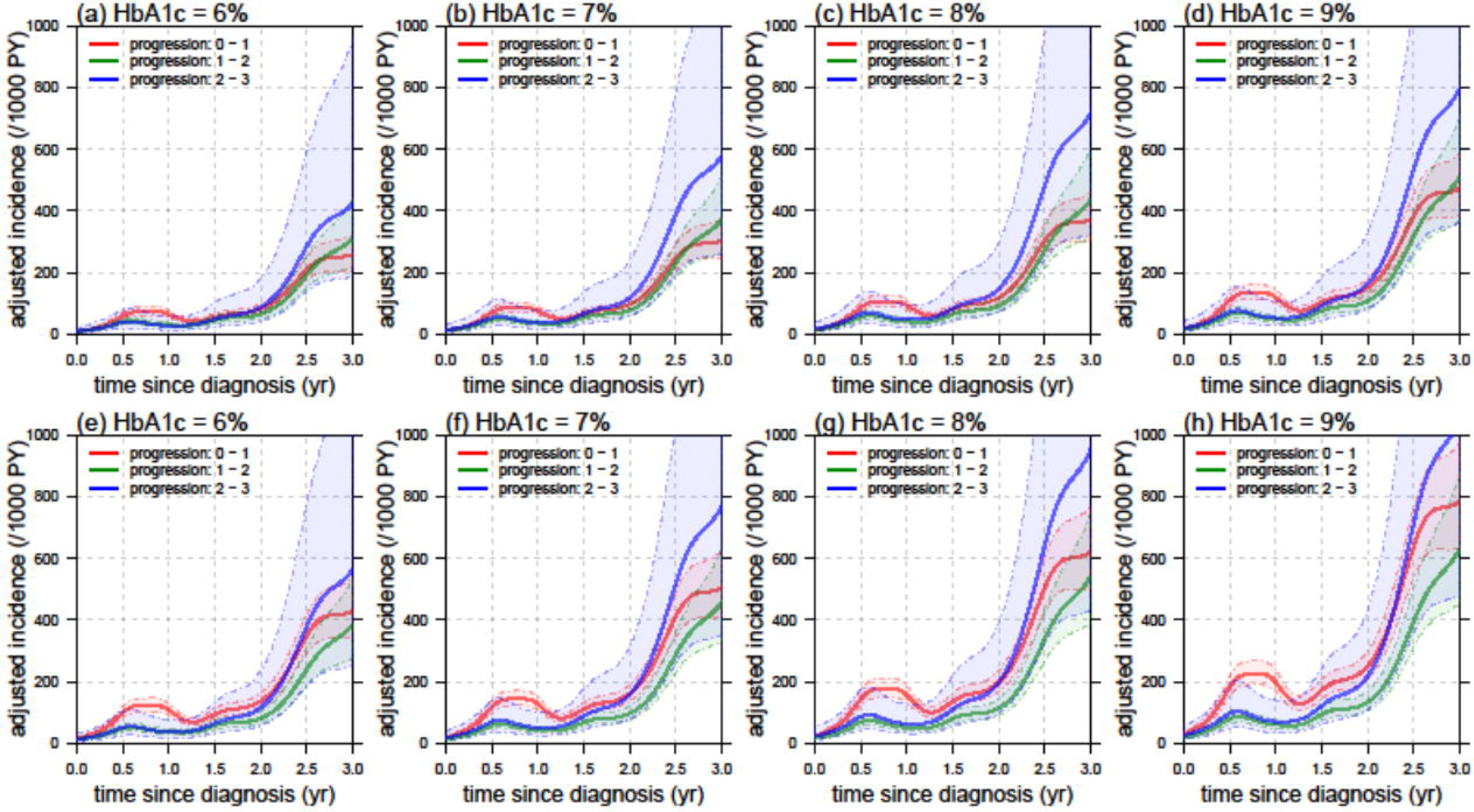
Adjusted incidence rates per 1,000 person-years (PY) of CKD new-onset or progression over time since the diagnosis of previous stages in female patients, at the HbA1c levels ranging from 6% to 9%, for new-onset (red solid line), progression from stage 1 to 2 (green solid line) and from stage 2 to 3 (blue solid line). Upper panel (a-d) show adjusted incidence rates in patients aged below 65 years; Lower panel (e-h) show adjusted incidence rates in patients aged 65 years or above. The broken line and shadow area indicate the 95% confidence interval.

**Figure 3.**
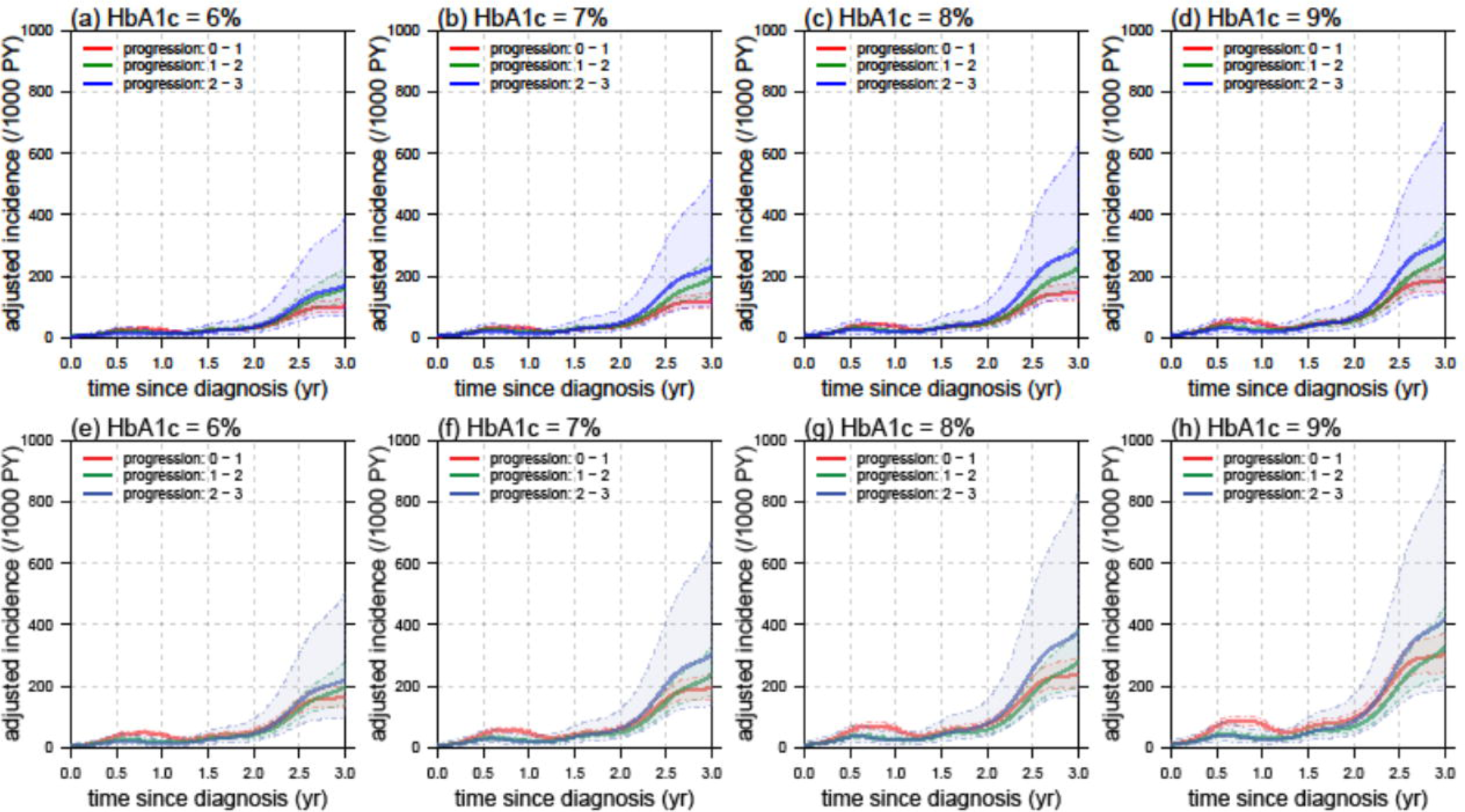
Adjusted incidence rates per 1,000 person-years (PY) of CKD new-onset or progression over time since the diagnosis of previous stages in male patients, at the HbA1c levels ranging from 6% to 9%, for new-onset (red solid line), progression from stage 1 to 2 (green solid line) and from stage 2 to 3 (blue solid line). Upper panel (a-d) show adjusted incidence rates in patients aged below 65 years; Lower panel (e-h) show adjusted incidence rates in patients aged 65 years or above. The broken line and shadow area indicate the 95% confidence interval.

### Rates of regression

The significant predictors for regression from stage 1 to 0 include male, younger age, lower levels of SBP and creatinine, without ACEI or ARB prescriptions (Appendix 3). The stage 2-1 and 3-2 regression has similar predictors, including male, older age, lower levels of SBP and creatinine. Higher levels of DBP and without ACEI are only significant for the stage 3-2 regression.

The regression from earlier stages shows a less evident temporal pattern in women and men (Appendix 4 and 5). The adjusted incidence rates were high during the first half year and then quickly declined to very low levels after one year. The regression rates were higher in the group from stage 2 to 1, followed by 1 to 0 and stage 3 to 2 in both sex groups. The regression rates of older people tend to be slightly higher than the younger group.

## Discussion

Early predictions of complication incidence and disease progression are of great concern in the clinical management of type 2 diabetes patients. The MSMS model has recently been introduced to evaluate the risk of chronic diseases ^21-23^. Its advantages over the classical PH model have been discussed in literature ^14 16 24^. This novel modeling approach allows the simultaneous prediction of multi-state transitions, either progression or regression, and thereby is capable of assessing both linear and nonlinear effects of predictors. Previous studies have demonstrated that the MSMS model outperformed the PH model in both model structure and fitting performance ^25-28^. Our study is among the first to apply this model to predict the risks of CKD progression and regression in a large sample of Chinese population with type 2 diabetes under primary care. The advantage of MSMS model also lies in its ability of incorporating nonlinear effects. We observed a rapidly increasing progression rates between different stages among female patients, but to a lesser extent among male patients (Figure 2 and 3). Particularly, a turning point of the progression rates was found at one year post diagnosis of previous stage, while the regression rates peaked at half a year. This suggests that one year is a critical control window to reverse the deterioration of nephropathy among patients with type 2 diabetes. Another critical time point is two years post diagnosis, beyond which the regression rates for both gender dramatically decreased, suggesting an irreversible trend of kidney function deterioration.

Based on the data availability and association with disease progression, we considered a series of predictors including demographic characteristics, clinical presentations, drug prescriptions, and laboratory tests routinely conducted in these patients. Interestingly, a larger number of significant predictors were successfully identified by the MSMS model for new-onset than for progression. Nearly all these predictors significantly predicted the CKD new-onset, whereas for progression, only age, gender, BMI, SBP and serum creatinine remained significant predictors. These findings highlight the importance of early interventions for type 2 diabetes patients when their renal functions maintain within the normal range and many risk factors are modifiable.

A few prediction models for progression of CKD have been developed in literature ^13 29 30^ and some were specifically for patients with diabetes ^31-33^. This study is among the first to define CKD progression (or regression) as a forward (or backward) transition between stages of proteinuria and deterioration of eGFR. In this study we found that female, older age, ACEI and ARB prescriptions, higher BMI, SBP, total cholesterol and creatinine, lower DBP, HDL and LDL were associated with a higher risk of CKD new-onset and progression. However, Tangri et al reported that younger age and male had a faster progression to renal failure in their PH model for CKD patients of various etiologies ^34^. This discrepancy might be due to the different outcome variables and study populations, since our cohort included only type 2 diabetes patients and the outcomes were progression from earlier stages. The predictors of our model are similar to an early prediction model that was developed using the PH model for cardiovascular diseases (CVD) of type 2 diabetes patients in Hong Kong ^35^. This CVD model found that older age, longer duration of type 2 diabetes and higher HbA1c predicted a higher risk of CVD incidence.

This study is among the first to investigate the predictors of CKD regression in type 2 diabetes patients under primary care, to our best knowledge. We found that younger age had a faster regression from stage 1 to 0, but slower from stage 2 to 1 or 3 to 2. This suggests that, once younger adults enter the advanced stages, it is less likely to reverse the progression.

There is ample evidence in literature to demonstrate that both ARB and ACEI could slow down the progression of CKD. However, we found that type 2 diabetes patients who were prescribed with ARB and ACEI had a dramatically faster progression rate of CKD and a lower regression rate. In our data, these drugs were significant only in transition of stages 0 to 2, and only ACEI prescription predicts a lower rate of regression from 3 to 2. We speculate that patients who are prescribed with ACEI and ARB more likely had a high-risk profile than did those without these prescriptions. Future research would be useful to investigate whether the association found in our study was truly due to the drugs themselves or confounding.

Previous studies including a few from Hong Kong have reported that men with type 2 diabetes had a higher risk of CKD incidence than women ^4^. On the contrary, we found that women had a faster rate of CKD progression, and men had a higher rate of regression in our cohort. Interestingly, we also found that female patients with different drug usage showed distinct temporal patterns of CKD progression, while less discrepancies were observed in the male groups. This gender heterogeneity has been well recognized in the development of diabetic complications, but not in drug efficacy. Further studies are warranted to explore this important gender heterogeneity.

The results of this study have important clinical implications. Few previous studies have considered the prediction model for progression of early stages, however, a timely intervention on these early stages in the primary care level could have a significant impact to prevent or slow down the further progression to the end stage renal failure. This would potentially reduce the disease burden and economic burden on the individual levels as well as on the secondary care of the health care system. Our prediction models were developed from the routine clinical data of electronic health records, thereby it could be automatically integrated into the existing clinical information system and easily adopted by clinicians to visualize the CKD progression risks in primary care settings. The graphic presentation of individualized CKD risk prediction could be a useful tool to facilitate the risk communication and health education for disease management of diabetes.

Interestingly, we found a small peak of CKD progression risks within half to one year since enrolment, even after we adjusted for check-up frequency (Figure 2 and 3). It is of note that these participants had different lengths of years with type 2 diabetes, and some already had developed complications when first enrolled into the RAMP-DM program. We speculate that these patients progressed faster than others but it was actually due to their delayed lab tests and clinical visits. Furthermore, although we adjusted the intervals of follow-up checks in the model by calculating adjusted incidence rates, it is difficult to completely eliminate the impact of inconsistent intervals of clinical visits and laboratory tests. Therefore, the peaks within one year need cautious interpretations. Ideally, the prediction risk curve should be plotted over the time since diagnosis. However, this could be very difficult to achieve, as many patients did not have timely laboratory tests for diagnosis. In fact, many prediction models also used the year since enrolment as time indicator, including a few local studies ^8 35^.

There are a few limitations in this study. Firstly, although we had a large sample size, the follow-up period was relatively short. CKD stages were defined by one test result due to the relatively short study period. Therefore, the predicted rates beyond two years tend to have wide confidence intervals. Secondly, a large numbers of patients in RAMP-DM were excluded due to incomplete eGFR, UACR and HbA1c data. It is unclear to us whether some were not scheduled to take tests due to mild conditions, or others skipped clinical visits for some reasons. Therefore, some of our model estimates might slightly overestimate or underestimate the true risks. Future studies with longer follow-up periods are warranted to obtain more accurate predictions. Last but not least, the study population were Chinese patients under primary care in a highly developed economic region. Future studies are warranted to apply our model to other populations as external validation and calibration, so that the models can eventually become a useful tool in clinical practice.

## Conclusion

The MSMS model achieved satisfactory performance in predicting the progression of CKD in patients with type 2 diabetes under primary care. The prediction model developed from this study could be applied to build an online calculator for individual risks of CKD progression. This will greatly facilitate clinical decision making of individualized intervention plan and treatment target to slow the progression of CKD in Chinese adults with type 2 diabetes, based on biomarkers of glycaemic control, cardiovascular and renal function. The online calculator will also improve the risk communications of doctors and nurses with patients.

## Data Availability

Data may be obtained from a third party and are not publicly available

## Ethical approval and informed consent

The ethical approval has been obtained from the New Territory West Cluster Clinical and Research Ethics Committee (NTWC/CREC/17085). Consent forms were exempted because all the anonymized data were extracted from the computerized data system of the Hospital Authority, and no personal information was collected for this study.

## Acknowledgements

We thank Nick Lau for assistance in data collection and Jay Hebert of The Hong Kong Polytechnic University for proofreading the manuscript.

## Accessibility of data and materials

Data may be obtained from a third party and are not publicly available

## Data sharing statement

No additional data available.

## Contributorship statement

LY, TKC designed the study and prepared the manuscript; Jun L, CWL and Jinxiao L contributed to data collection and result interpretation; SZ, DH and JQ conducted the data analysis; all authors have reviewed and proved the final version of this manuscript for publication.

